# ΦB124-14 *Bacteroides* Bacteriophage-Like Quantitative Real-Time PCR Assays for Human Sewage Pollution Measurement

**DOI:** 10.1101/2025.04.23.25326295

**Authors:** Jessica R. Willis, Mano Sivaganesan, Brian McMinn, Asja Korajkic, Christopher Staley, Orin C. Shanks

## Abstract

The ability to measure human sewage in environmental and wastewater samples is important to protect public health and natural water resources. A recent study reports the complete genome sequence of ΦB124-14, a bacteriophage capable of infecting a narrow subset of *Bacteroides* spp. closely associated with the human gut. To investigate the use of ΦB124-14 for fecal indicator and source identification applications, the entire genome was interrogated for potential human-associated genetic regions using a combination of bioinformatic and laboratory testing approaches. To assess bioinformatic predictions, 53 primer sets were subject to systematic testing using 100 fecal samples from ten animal sources, primary influent sewage from 36 geographical locations across the continental United States, and environmental surface waters with known human sewage impact. Based on candidate primer set end-point PCR analyses and next generation amplicon sequencing, two novel hydrolysis probe-based quantitative real-time PCR assays (qPCR), PS28 and PS30, were designed and evaluated. Both qPCR assays exhibited a sensitivity of 86.1%, a specificity of 100%, and successfully detected ΦB124-14-like genetic markers in sewage impacted environmental water samples. PS28 and PS30 performance was then compared to top performing DNA-based viral (CPQ_056 and CPQ_064) and bacterial (HF183/BacR287 and HumM2) human-associated qPCR assays. Findings indicate ΦB124-14 bacteriophage-like qPCR assays exhibit superior specificity, but markers consistently occur at lower concentrations in United States primary influent sewage samples. Furthermore, paired measurements of ΦB124-14 and crAssphage bacteriophage-like sequences in high volume (10 L) primary influent sewage samples (*n* = 38) indicate significant correlations ranging from r = 0.593 (p < 0.0001; PS30 versus CPQ_056) to r = 0.938 (p < 0.0001; PS28 versus PS30). A comparison of bacteriophage-like marker concentrations with cultured GB-124 in sewage samples showed no significant correlations (r ≤ 0.215, p ≥ 0.183). Finally, novel qPCR methods for potential human sewage management applications and future research directions are discussed.

## 1. INTRODUCTION

The presence of human sewage in environmental waters remains a key public health concern due to the potential for harboring disease-causing microorganisms. As a result, sewage is often treated prior to discharge into environmental receiving waters. Most public health managers rely on bacterial pathogen surrogates or general fecal indicator bacteria (i.e., *Escherichia coli* and enterococci) to evaluate the efficacy of wastewater treatment and to detect sewage pollution in surface waters. However, general bacterial fecal indicators may not always respond to wastewater treatment or environmental stressors (i.e., ultraviolet radiation) in a similar manner to pathogens, especially enteric viruses such as noroviruses, enteroviruses, and adenoviruses which are often responsible for waterborne disease [1–7]. Another limitation of general fecal indicator bacteria methods is their incapacity to discriminate between human sewage and other potential sources, preventing focused remediation because many waterbodies are polluted by a combination of human sewage and feces from companion animals (i.e., dogs) and/or wildlife sources. These disadvantages have led to the development of numerous virus-based methodologies targeting crAssphage and other *Bacteroides* bacteriophages [8–10], polyomaviruses BK and JC [11], pepper mild mottle viruses [12], and tomato brown rugose fruit viruses [13], among others. Most of these methods rely on the detection of nucleic acid targets (i.e., DNA or RNA) rather than cultivation, often limiting the ability to confirm the presence of infectious viruses in a sample.

A promising cultivation-based method targets a bacteriophage that infects *Bacteroides fragilis* strain GB-124 [8]. The GB-124 method is reported to exhibit strong human fecal association [8, 14], occur in wastewater across multiple geographic regions [14–17], co-occur with norovirus and adenovirus in wastewater [18], exhibit a similar response to ultraviolet radiation inactivation with coxsackievirus, echovirus, hepatitis A virus, poliovirus, and rotavirus [19], and has been detected in sewage polluted surface and reclaimed waters [17]. However, an ideal viral human-associated indicator toolbox should contain methodologies allowing for both cultivation and genetic testing.

A recent study reports the complete genome sequence of ΦB124-14, a bacteriophage capable of infecting a narrow subset of *Bacteroides* spp. including the *B. fragilis* GB-124 strain and ecological profiling suggests it is closely associated with the human gut [20]. The double stranded DNA ΦB124-14 genome is 47,159 base pairs in length and encodes for 68 predicted open reading frames (ORF) with non-coding sequence regions limited to 8.2% of the genome. A subsequent study determined that an ΦB124-14 ecogenomic signature was able to delineate between human and non-human fecal metagenomic DNA sequencing libraries [21]. Furthermore, it was detected in environmental datasets with known human fecal pollution, prompting the identification of multiple genetic regions predicted to exhibit a strong human fecal association [21]. However, attempts to develop a PCR-based ΦB124-14 bacteriophage-like assay have been unsuccessful.

Here, we employed a “biased genome shotgun strategy” used to develop crAssphage-like qPCR assays [9] to interrogate the ΦB124-14 genome for human sewage-associated genetic regions. Select genetic regions of the ΦB124-14 genome were then screened using end-point PCR for highly specific and abundant sewage-associated DNA targets. These DNA targets were subsequently used to develop two novel qPCR assays as potential future wastewater and surface water testing tools. Novel ΦB124-14 bacteriophage-like qPCR assay performance was then compared with top performing DNA-based bacterial (HF183/BacR287 and HumM2) and viral (CPQ_056 and CPQ_064) human-associated methodologies. Finally, potential trends between primary sewage influent paired measurements of ΦB124-14 and crAssphage bacteriophage-like qPCR assays, as well as culture-based GB-124 were investigated. Findings suggest that bioinformatic predictions combined with high-throughput laboratory screening of the ΦB124-14 genome successfully led to the development of sewage-associated qPCR methods that may be important for future research and wastewater management activities.

## 2. MATERIALS & METHODS

### 2.1 Sample collection

Individual fecal samples (*n* = 100) were collected from different locations across the United States, as previously described [22]. Fecal samples represent nine species including *Anser* spp. (Canada goose; *n* = 10), *Canis familiaris* (dog, *n* = 14), *Bos taurus* (cow, *n* = 10 and calf, *n* = 10), *Equus caballus* (horse, *n* = 10), *Gallus gallus* (chicken, *n* = 10), *Sus scrofa* (pig, *n* = 10), *Procyon lotor* (raccoon, *n* = 10), and *Halichoerus grypus* (grey seal, *n* = 10) (see **Table S1** for sample details). Each sample was collected from a different individual and stored at -80℃ until time of DNA extraction (< 18 months).

The collection and handling of grey seal samples was conducted under the authority and in strict accordance with the National Oceanographic and Atmospheric Administration Marine Mammal Protection Act (Permits: 17670A and 21719). Single grab primary influent sewage was collected from 36 facilities situated across the contiguous United States as previously described [16, 22]. Briefly, 1 liter of untreated primary influent was collected, immediately stored on ice, and shipped overnight for laboratory testing (maximum holding time = 24 h). Study participation was voluntary. Finally, as a proof-of-concept, surface water samples were collected from the Heiserman Stream (East Fork Watershed, southwest OH, USA) downstream of a wastewater treatment facility discharge outfall. These samples were collected in sterile 1 L containers, immediately stored on ice, and transported to the laboratory for DNA extraction testing (< 4 h).

### 2.2 DNA extraction and quantification

DNA was extracted from 10 mL of primary influent sewage as previously described [16] using a modified QIAamp Blood Maxi Kit protocol (Qiagen, Valencia, CA, USA). For fecal samples, DNA was extracted using the DNeasy PowerSoil Pro Kit (Qiagen) according to manufacturer’s instructions with the following modifications. Approximately 0.25 g of fecal material was used. Cell lysis was achieved with a MP FastPrep-24 (MP Biomedicals, LLC Solon, OH, USA) at 6.0 m/s for 30 s. DNA was eluted with 100 µL TE buffer (Tris-HCl 10 mM, EDTA 0.1 mM, pH 8.0). Sewage and fecal DNA extract concentrations were determined with a Qubit 3 Fluorometer (Thermo Fisher Scientific, Grand Island, NY, USA), diluted to 0.5 ng/µL to normalize sample test quantities for performance testing, and stored in low-retention microcentrifuge tubes at -20°C until PCR-based testing (< 18 months). For each environmental water sample, 200 mL was concentrated to a final volume of approximately 200 µL using an automated Concentrating Pipette with a single-use ultrafiltration hollow fiber polysulfone tip following the manufacturer’s instructions recommended for virus concentration (InnovaPrep, Drexel, MO, USA). The entire elution volume was subject to DNA extraction using the AllPrep PowerViral DNA/RNA Kit manufacturer’s protocol with the omission of the bead milling step and use of 100 µL of TE buffer for elution. Water sample DNA extracts were stored at 4°C in low-retention microcentrifuge tubes until the time of amplification (< 24 h). For each batch of DNA extractions, three method extraction blanks were performed to monitor for potential contamination by substituting purified water for fecal, sewage, or environmental water samples.

### 2.3 Selection of candidate genetic regions for PCR-based method development

To identify candidate genetic regions, select portions of the approximately 47.2 kbp ΦB124-14 genome (accession code: HE608841.1) [20] were identified for end-point PCR testing using a two-step process. First, to select for candidate regions with some evidence of genetic conservation, only ΦB124-14 genome predicted coding regions (*n* = 68; length range: 114 to 6,021 bp) were considered due to the potential for rapid DNA mutation rates in the human gut virome [23]. Second, BLASTn was performed for each open reading frame (ORF) coding sequence using the nr database (performed 4/5/2020). Any DNA sequence > 100 bp with either no significant hit (E-value > 0.001) or significant hit with a sequence identity < 70% compared to the *Bacteroides* phage B40-8 reference genome (accession code: FJ008913.1) [24] was considered a candidate genetic region for end-point PCR primer set development. Genetic regions with sequence identities ≥ 70% compared to the *Bacteroides* phage B40-8 reference genome were excluded to further minimize selection of DNA sequences that may be conserved between closely related bacteriophage that infect *Bacteroides fragilis* because 16S rRNA genetic markers targeting this species have been detected in non-human fecal materials [25]. At the time of bioinformatic assessment, the *Bacteroides* phage B40-8 genome was the most closely related to ΦB124-14 (57% sequence identity) [20].

### 2.4 Candidate primer design

A total of 53 end-point PCR primer pairs were designed to amplify selected ΦB124-14 regions. Primer pairs were designed and tested *in silico* using Primer-BLAST [26] with default parameters, except that product length was constrained to 80-160 bp. Primer pair specificity was evaluated using the refseq representative genomes bacterial virus (taxid: 28883) database (4/6/2020), and primer specificity stringency was set to: primer must have ≥ 1 mismatch to unintended targets, ≥ 1 mismatch within the last five bp at the 3’ end, and ignore targets with ≥ 2 mismatches to primer. Only primer pairs that generated BLAST hits (E-value < 0.001) to ΦB124-14 sequences were selected. Genetic regions where no primer sets met design criteria were eliminated from further consideration.

### 2.5 End-point PCR amplification and gel visualization

Each 25 µL end-point PCR amplification consisted of TaKaRa Ex *Taq* DNA Polymerase Hot-Start PCR reagents (Takara Bio USA, San Jose, CA, USA), 200 nM each of the forward and reverse primers, 4% bovine serum albumin (BSA; Thermo Fisher Scientific), 2 µL of template DNA, and molecular grade water. End-point PCR tests were performed in triplicate on a MiniAmp Thermal Cycler (Thermo Fisher Scientific) under the following conditions: 94°C for 5 m followed by 40 cycles of 94°C for 40 s, 60°C for 1 m, and 72°C for 30 s. During end-point PCR amplification, a minimum of two no-template controls (NTC) containing purified water instead of template DNA were performed with each instrument run to monitor for potential sources of extraneous DNA. PCR product results were visualized via capillary electrophoresis with the QIAxcel Advanced instrument using a DNA Screening Kit (Qiagen) according to manufacturer’s instructions.

### 2.6 Candidate end-point PCR primer set evaluation

To determine which candidate ΦB124-14 bacteriophage-like genetic regions have human fecal source identification potential, each primer set was tested with end-point PCR in a three-round process. In the first round, candidate primer sets were challenged against two fecal DNA composites representing sewage and non-human sources. The sewage DNA composite (1 ng of DNA per reaction) consisted of equal amounts of primary influent sewage DNA from five geographic locales (0.2 ng of DNA per reaction). The non-human DNA composite (4 ng of total DNA per reaction) consisted of equal amounts of DNA (0.2 ng of DNA per reaction per animal group) from five groups including pig (*n* = 5), cow (*n* = 5), dog (*n* = 5), and raccoon (*n* = 5) fecal DNA. Candidate primer sets proceeded to a second round of testing if the following criteria were met: (1) a PCR product of expected size was present when the primary influent sewage DNA composite was used as a template, (2) a PCR product of expected size was absent when the non-human DNA composite was used as a template, (3) a PCR product was absent when purified water was used as a template, and (4) absence of non-specific PCR product sizes, excluding primer-dimers.

In round two, the remaining candidate primer sets were challenged against diluted preparations of the primary influent sewage DNA composite (0.1, 1 × 10^-2^, and 1 × 10^-3^ ng per reaction) and a higher concentration (5 ng of total DNA per reaction) of separate non-human species composites for pig (*n* = 10), cow (*n* = 10), dog (*n* = 10), and raccoon (*n* = 10). Candidate primer sets proceeded to a third round of testing under the following conditions: (1) amplification of expected size when the primary influent sewage composite was used as a template for one or more test concentrations, (2) absence of expected PCR product in all reactions when a non-human DNA composite was used as the template, (3) absence of amplification when purified water was used as a template, and (4) absence of non-specific PCR product sizes, excluding primer-dimers.

Round three represented the most rigorous performance screening step for candidate end-point PCR primer sets. Evaluation began with specificity determination based individual testing of 100 fecal samples (1 ng of DNA per reaction), followed by geographic distribution characterization in primary influent sewage samples collected from 36 locations (1 ng of DNA per reaction). If a candidate primer set achieved a specificity ≥ 95%, then it was subjected to sewage geographic distribution and environmental water sample testing. Top-performing primer sets were defined as exhibiting ≥ 80% specificity and ≥ 80% sensitivity [27].

### 2.7 DNA sequence verification of top-performing end-point PCR primer sets

To verify that candidate primer sets passing round three screening were amplifying the intended ΦB124-14 genetic region, amplification products from four primary influent sewage samples were sequenced and evaluated. End-point PCR was performed using primer pairs that passed round three, using the same amplification conditions as above. Amplicons were paired-end sequenced at a read length of 301 nucleotides (nt) on the Illumina MiSeq platform by the University of Minnesota Genomics Center. Raw reads are deposited in the NCBI SRA under BioProject accession <u>SRP576867</u>. Sequence processing was done using mothur software version 1.48.1 [28]. Forward and reverse reads were trimmed to 70 and 90 nt for primer set 28 and 30 amplicon libraries, respectively, and pair-end joined using fastq-join software [29]. Joined sequences were screened for primer sequences, which were subsequently removed. Amplicon sequence variants (ASV) were calculated using the furthest-neighbor algorithm at distance equals 0.10 sequence similarity (reported at ≥ 100× coverage).

### 2.8 ΦB124-14 Bacteroides bacteriophage-like qPCR assay design

Candidate primer sets passing round three end-point PCR testing were adapted to hydrolysis probe-based qPCR chemistry. Primers and probes for putative human-associated ΦB124-14-like genetic regions were designed using default parameters of the Primer Express software (version 3.0.1; Thermo Fisher Scientific). Fluorogenic minor groove binding (MGB) probes were 5’ labeled with 6-carboxyfluoroscein (FAM).

### 2.9 qPCR standard DNA material preparation

Standard DNA material sources consisted of three plasmid constructs. The commercially available National Institute of Standards and Technology Standard Reference Material^®^ 2917 (SRM 2917; Rockville, MD) and a customized ΦB124-14 construct (Integrated DNA Technologies, Coralville, IA) were used for qPCR calibration model generation. An internal amplification control (IAC; Integrated DNA Technologies) was prepared for amplification inhibition screening using the duplex HF183/BacR287 qPCR assay [30]. Both the ΦB124-14 plasmid construct and IAC were linearized by ScaI-HF restriction digest (New England BioLabs, Beverly, MA), purified via QIAquick PCR Purification Kit (Qiagen), quantified with a Qubit dsDNA HS assay kit on a Qubit 3 Fluorometer (Thermo Fisher Scientific), and diluted in 10 mM Tris-Cl 0.5 mM EDTA (pH 9.0). For the ΦB124-14 standard material, five dilutions were prepared to contain 10 to 1 × 10^5^ copies per 2 µL. The IAC material was diluted to generate 10^2^ copies/2 µL. All DNA material preparations were stored in low-retention microtubes either at 4°C (SRM 2917) or -20°C (ΦB124-14 and IAC).

### 2.10 qPCR amplifications

A total of six qPCR assays were used in this study: two novel ΦB124-14-like assays (this study) and four previously reported DNA-based human-associated fecal source identification methods [9, 30, 31] including two bacterial assays (HF183/BacR287 and HumM2) and two viral assays (CPQ_056 and CPQ_064). All reactions contained 1× TaqMan Environmental Master Mix (version 2.0; Thermo Fisher Scientific), 0.2 mg/mL bovine serum albumin (Sigma-Aldrich), 1 µM of each primer, 80 nM FAM-labeled probe, 80 nM VIC-labeled probe (HF183/BacR287 assay only), and molecular grade water. The total volume of each reaction was 25 µL containing 2 µL of template (sample DNA, reference material, or molecular grade water). HF183/BacR287 duplex reactions also contained 10^2^ copies of IAC template. All qPCR test plates included a standard curve and reactions were performed in triplicate using the QuantStudio 3 PCR System (Thermo Fisher Scientific) with the following thermal cycle profile: 10 m at 95°C followed by 40 cycles of 15 s at 95°C and 1 m at 60°C. Fluorescence thresholds were manually set to either 0.08 ΔRN (HumM2) or 0.03 ΔRN (other assays). Quantification cycle (C_q_) values were exported to Microsoft Excel for further analysis. To monitor for potential extraneous DNA contamination, six NTC reactions with molecular grade water substituted for template DNA were performed with each instrument run.

### 2.11 Performance comparison testing of ΦB124-14-like bacteriophage-like qPCR assays

To investigate the suitability of newly developed technologies for human fecal source identification applications, the performance of ΦB124-14-like qPCR methods was evaluated in a series of experiments and compared to four established human-associated qPCR assays (HF183/BacR287, HumM2, CPQ_056, and CPQ_064). All fecal source identification qPCR assays were subject to calibration model acceptance criteria for linearity (R^2^ ≥ 0.980) and amplification efficiency (*E*; 0.90 to 1.10) [32, 33]. Next, the abundance of each genetic marker was measured in primary influent sewage samples (*n* = 36) at a test concentration of 1 ng of DNA per reaction. The prevalence of human-associated genetic markers in non-human pollution sources was evaluated by testing 100 individual non-human fecal samples from 10 animal groups at a test quantity of 1 ng of total DNA per reaction. Finally, as a proof-of-concept pilot demonstration, genetic marker concentrations were estimated from three environmental water samples (2 µL of sample DNA extract per reaction).

### 2.12 Comparison of culture-based GB-124 with ΦB124-14 and crAssphage bacteriophage-like qPCR genetic targets

To explore potential associations between novel ΦB124-14 and crAssphage bacteriophage-like qPCR targets and cultivated levels of GB-124 plaque forming units (PFU), data and sample material from 38 primary influent sewage samples collected from two wastewater treatment facilities in a previous study were used [34]. For *B. fragilis* GB-124 bacteriophage measurements, previously reported concentrations (PFU/L) were used from primary influent sewage samples (10 L) concentrated by dead-end hollow-fiber ultrafiltration (D-HFUF; [35]) followed by a modified single agar layer method [34, 36]. ΦB124-14 and crAssphage bacteriophage-like qPCR target concentrations were measured from DNA extractions (this study) of archived celite concentrates (previous study [34]). Briefly, celite pelleted viral nucleic acid material from 200 mL of D-HFUF concentrate was resuspended in 20 ml of phosphate buffered saline (pH 9.0). Then, nucleic acids from 200 µL of resuspension was extracted using the Qiagen AllPrep PowerViral Kit (Qiagen) according to manufacturer’s protocol. Two microliters of resulting DNA extract were used as template for each qPCR reaction, equivalent to 2 mL of the original primary influent sewage sample. CPQ_056 was used to estimate crAssphage-like sequences due to a strong correlation with CPQ_064 in untreated sewage samples from 36 geographic locations observed in this study (r = 0.988; p < 0.0001).

### 2.13 Data analysis and amplification inhibition monitoring

‘Single’ calibration models were generated for each qPCR assay and instrument run combination using a Bayesian Markov Chain Monte Carlo approach [37, 38]. Outliers were defined as the absolute value of the studentized residual > 3. Amplification efficiencies (*E*) were based on the following equation: *E* = 10^(-1/slope)^ – 1. The lower limit of quantification (LLOQ) was defined as the instrument run specific 95% Bayesian credible interval upper-bound from repeated measures of the respective reference DNA material lowest concentration dilution.

HF183/BacR287 duplex IAC protocol was used to monitor for amplification inhibition in all DNA extracts [39, 40]. Any DNA extract with evidence of amplification inhibition was discarded from the study. Instrument run-specific IAC proficiency testing (HF183/BacR287 NTC VIC C_q_ standard deviation ≤ 1.16) was also included to confirm reliable application of amplification inhibition monitoring from one instrument run to another [39, 40]. Specificity was defined as TNC/(TNC + TPI), where TNC represents the total number of negative samples that tested negative correctly (negative result = non-detection for all sample replicates), and TPI is the total number of samples that tested positive incorrectly (positive result = detection in one or more sample replicates). Pearson product momentum correlation analysis was performed between paired measurements (α = 0.05). All statistics were conducted with SAS software (Version 9.4 M8; Cary, North Carolina, USA) or WinBugs (Version 1.4.3; https://www.mrc-bsu.cam.ac.uk/software/bugs/).

## 3. RESULTS

### 3.1 Putative human-associated ΦB124-14 bacteriophage-like genetic regions and candidate primer set design

A total of 12,026 bp (25.6%) of the ΦB124-14 genome were selected for end-point PCR screening to identify potential human-associated genetic regions. ΦB124-14 select genetic regions were excluded due to noncoding assignments (8.2%) and similarities to the *Bacteroides* phage B40-8 genome (66.2%). Of the 12,026 bp selected for end-point PCR testing, 5,795 bp (48.2%) were not amenable for PCR testing based on primer design parameters (e.g., product size and melting temperature restrictions). In total, 53 end-point PCR primer sets were designed with 51.8% coverage (6,231 bp) of select putative human-associated genetic regions (**Table S1**).

### 3.2 Identification of human-associated ΦB124-14 bacteriophage-like genetic regions with end-point PCR

Candidate primer sets were subjected to three rounds of performance testing to identify putative human-associated ΦB124-14 bacteriophage-like genetic regions. In the first round, primers were tested against a sewage DNA composite and a non-human fecal DNA composite. Round one testing eliminated 46 (86.8%) candidate primer sets: 28.3% (*n* = 15) failed to yield a clearly distinct PCR product of the expected size in the sewage composite, 81.1% (*n* = 43) generated spurious PCR products (excluding primer dimerization byproducts), and 22.6% (*n* = 12) yielded false-positive results in non-human composite tests. Twelve (22.6%) candidate primer sets failed more than one criterion. Seven candidate primer sets were deemed eligible to move forward to round two testing (PS5, PS6, PS28, PS30, PS36, PS37, and PS43).

In round two testing, primer sets were challenged against dilutions of the sewage composite DNA and higher concentrations of separate non-human fecal composites representing four animal groups. Five primer sets were eliminated due to failing one or more criteria including presence of amplification product of expected size when a non-human sample was used as a DNA template (*n* = 2), failure to consistently yield a PCR product of the expected size when 0.1 ng of total sewage composite DNA per reaction was used as template (*n* = 1), or there was evidence of spurious PCR byproducts, excluding primer dimerization (*n* = 5). With each non-human group tested there was one false positive to pig and one false positive to dog observed. Two primer sets passed round two testing, including PS28 and PS30.

Round three testing included specificity determination with an expanded non-human fecal material collection and characterization of geographic distribution in sewage. PS28 exhibited the best performance with 100% specificity and 94.4% sensitivity to sewage samples. PS30 displayed 97% specificity (three false positives; 1 of 10 for chicken, cat, and calf samples) and 83.3% sensitivity. Please refer to Supplemental Information **Table S2** for detailed information.

### 3.3 DNA sequencing verification

End-point PCR products from PS28 and PS30 primer sets were sequenced from primary influent sewage samples to confirm amplification of expected ΦB124-14 bacteriophage-like sequences. Sequencing resulted in 402,748 raw sequencing reads (PS28 = 205,984; PS30 = 196,764) with 354,768 reads available after processing (PS28 = 169,850; PS30 = 184,918). The number of processed reads ranged from 38,925 to 50,841 across site and end-point PCR primer set combinations. A total of 16 amplicon sequence variants (ASV) were identified(**Figure 1**). PS30 sequencing indicated a single ASV (PS30_ASV_1) with 100% sequence identity to the ΦB124-14 genetic region, making up 99.9% of the processed sequence reads. In contrast, PS28 analysis yielded 16 ASVs with the three most abundant sequences accounting for 92.7% of processed sequence reads. PS28-ASV_7 (0.6% of PS28 processed sequence reads) exhibited the closest sequence identify to the expected ΦB124-14 genetic region with three mismatches.

**Figure 1:**
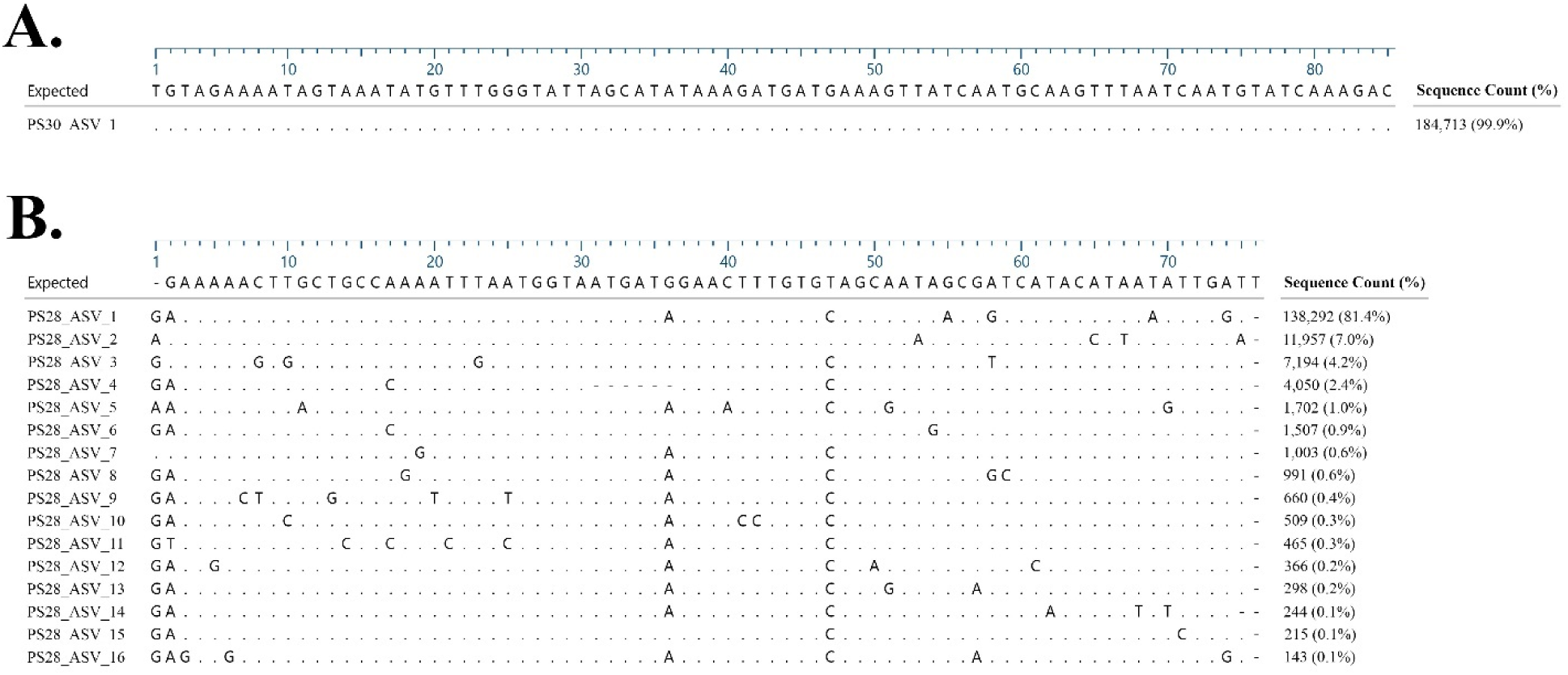
Alignment of sequences isolated from four primary influent sewage samples for PS30 (Panel A) and PS28 (Panel B) end-point PCR assays. ASV denotes amplicon sequence variant. The total number of observed sequences for each ASV is shown with the corresponding percent of total sequences in parentheses.

### 3.4 Performance of ΦB124-14 bacteriophage-like qPCR assays

Systematic testing of 53 primer sets identified two genetic regions (PS28 and PS30) for qPCR method development. Sequence information for primers and probes is listed in **Table 1**. The performance of these assays was evaluated through a series of paired experiments with established HF183/BacR287, HumM2, CPQ_056, and CPQ_064 protocols. The ΦB124-14 bacteriophage-like qPCR assays exhibited excellent calibration model performance, comparable to measurements from established assays (**Table 2**). Calibration R^2^ values were ≥ 0.997 and *E* values ranged from 0.90 to 1.03. All assays exhibited a range of quantification from approximately 10 to 10^5^ (ΦB124-14 target plasmid used for new PS28 and PS30 qPCR assays) or 10.3 to 1.04 × 10^5^ (SRM 2917 used for HF183/BacR287, HumM2, CPQ_056, and CPQ_064 qPCR assays) copies per reaction (full range of tested concentrations).

**Table 1:**
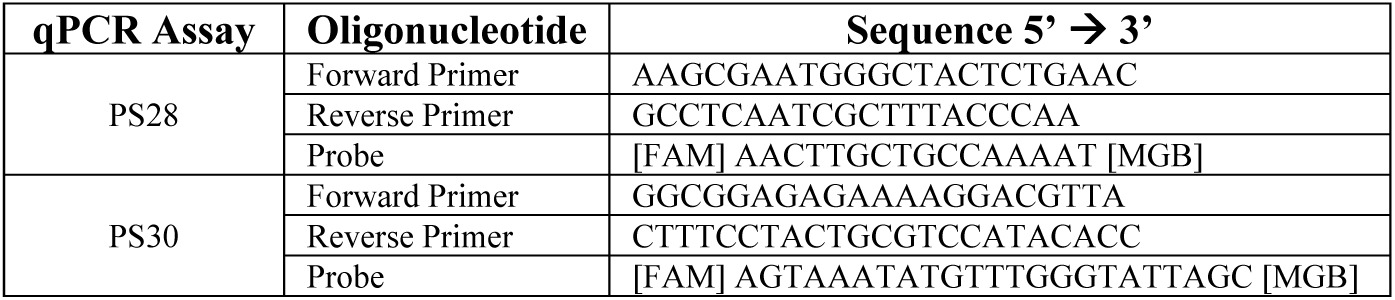
ΦB124-14 bacteriophage-like qPCR assay oligonucleotides.

**Table 2:** Summary of ΦB124-14 bacteriophage-like and established human-associated qPCR calibration model performance metrics.

| Assay | n | R<sup>2</sup> | Slope | Intercept | E | LLOQ |
| --- | --- | --- | --- | --- | --- | --- |
| PS28 | 9 | 0.997 to 0.999 | -3.46±0.03 to -3.29±0.04 | 37.5±0.10 to 38.3±0.10 | 0.95 to 1.02 | 34.5 to 35.2 |
| PS30 | 9 | 0.997 to 0.999 | -3.50±0.03 to -3.25±0.02 | 38.1±0.07 to 39.1±0.12 | 0.93 to 1.03 | 35.1 to 36.0 |
| CPQ_056 | 9 | 0.997 to 0.999 | -3.53±0.06 to -3.32±0.05 | 38.3±0.16 to 39.1±0.21 | 0.92 to 1.00 | 35.2 to 36.1 |
| CPQ_064 | 7 | 0.999 to 0.999 | -3.59±0.03 to -3.36±0.04 | 40.0±0.12 to 41.0±0.10 | 0.90 to 0.98 | 36.9 to 37.6 |
| HF183/BacR287 | 9 | 0.998 to 0.999 | -3.45±0.03 to -3.35±0.03 | 36.1±0.10 to 36.6±0.11 | 0.95 to 0.99 | 33.0 to 33.4 |
| HumM2 | 6 | 0.998 to 0.999 | -3.50±0.04 to -3.33±0.02 | 38.6±0.07 to 39.3±0.14 | 0.93 to 1.00 | 35.4 to 36.1 |

'n' shows the number of instrument runs for each qPCR assay.
R<sup>2</sup> signifies linearity of the fitted curve.
E indicates amplification efficiency [ $10^{(-1/\text{slope})} - 1$ ].
LLOQ represents lower limit of quantification reported as the quantitative threshold ( $C_q$ ).

Findings indicate ΦB124-14 bacteriophage-like qPCR assays exhibit 100% specificity based on systematic testing of 100 individual non-human fecal samples from 10 animal groups. In comparison, established human-associated qPCR assays yielded specificities ranging from 92% (CPQ_056) to 98% (HF183/BacR287). HF183/BacR287 (98% specificity) cross-reacted with two dog samples below the assay LLOQ (false positives ≥ 38.7 C_q_). HumM2 (95% specificity) cross-reacted with three calf (false positives ≥ 37.1 C_q_) and two cat (false positives ≥ 35.9 C_q_) samples, all below the assay LLOQ (**Table 2**). CrAssphage bacteriophage-like qPCR assays (specificity 92% for CPQ_056 and 95% for CPQ_064) cross-reacted with four cat samples within the range of quantification (range: 2.74 ± 0.001 to 3.08 ± 0.001 log_10_ copies per ng of DNA) as well as two additional cat samples (CPQ_056 only, ≥ 37.6 C_q_), a chicken sample (both assays, ≥ 36.4 C_q_), and a raccoon sample (CPQ_056 only, single replicate = 38.1 C_q_), all below the respective assay LLOQ (**Table 2**).

ΦB124-14 bacteriophage-like qPCR assays yielded 86.1% sensitivity to 36 geographically dispersed primary influent sewage samples (test quantity = 1 ng of DNA per reaction). Established human-associated qPCR assays exhibited 100%, consistently occurring at higher concentrations compared to PS28 and PS30 (**Figure 2**). While PS28 and PS30 were detected in most sewage samples, only four samples yielded log_10_ copies per ng of DNA concentrations within the range of quantification (range: 0.91 ± 0.03 to 1.02 ± 0.03). In contrast, established human-associated qPCR assays yielded measurements in the range of quantification for all samples with mean log_10_ copies per ng of DNA including 3.77 ± 0.33 (CPQ_064), 3.73 ± 0.34 (CPQ_056), 3.13 ± 0.39 (HF183/BacR287), and 2.15 ± 0.38 (HumM2). CrAssphage bacteriophage-like target sequence concentrations (log_10_ copies per ng of DNA) were significantly higher compared to HF183/BacR287 and HumM2 data sets (p < 0.001), but not between each other (p = 0.689). HF183/BacR287 measurements were significantly higher than HumM2 (p < 0.001). Proof-of-concept surface water samples impacted by treated effluent sewage exhibited a similar trend where HF183/BacR287, CPQ_056, and CPQ_064 yielded quantifiable levels in all water samples ranging from 2.39 ± 0.02 log_10_ copies per reaction (CPQ_064) to 2.60 ± 0.01 log_10_ copies per reaction (HF183/BacR287). However, HumM2 and PS30 testing resulted in detections below respective assay LLOQ for all three samples, while PS28 was only detected in one sample.

**Figure 2:**
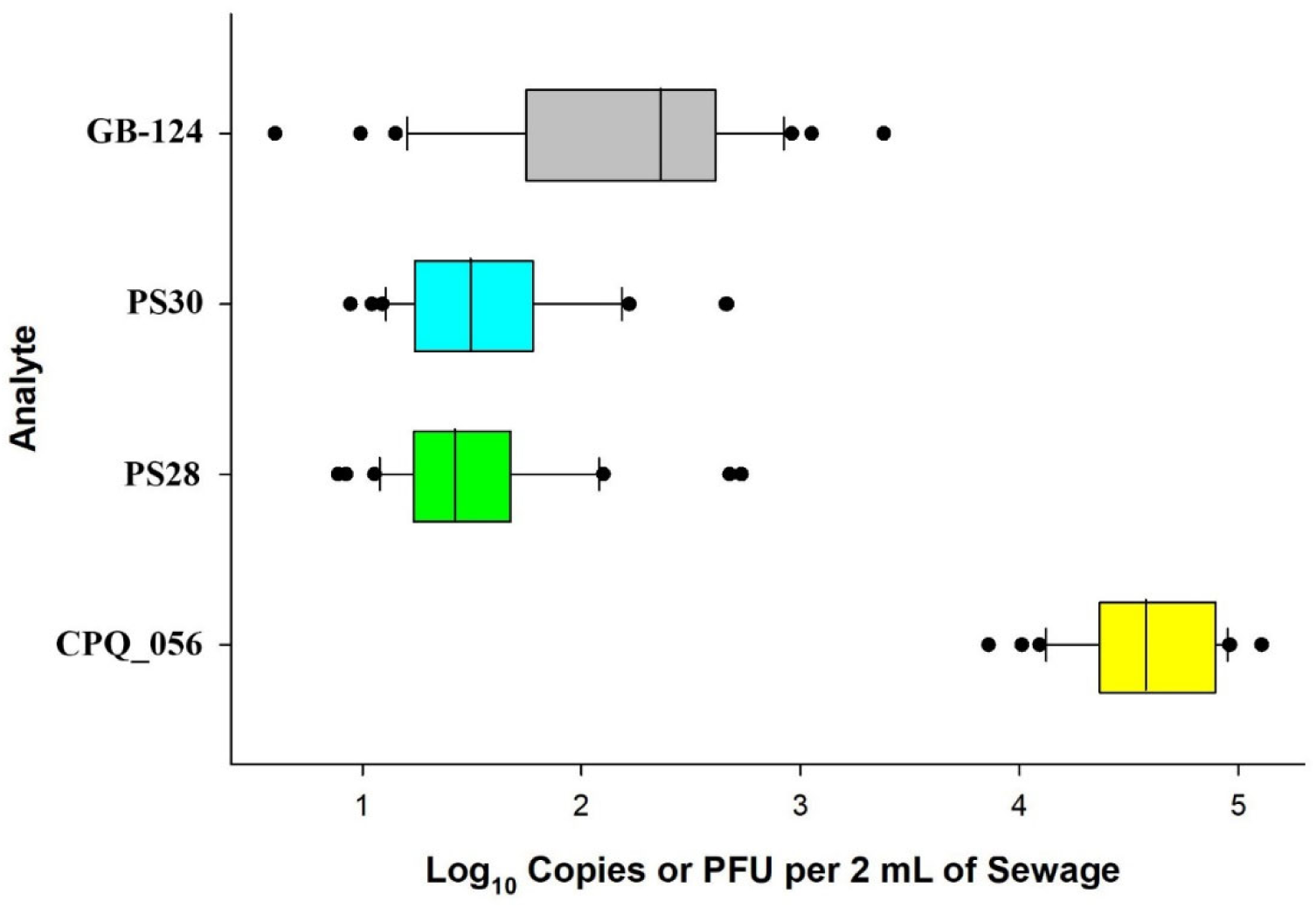
Abundance of ΦB124-14 and crAssphage bacteriophage-like target sequences (log_10_ copies estimates per 2 mL of sewage) and previously reported GB-124 cultivation paired measurements (PFU per 2 mL of sewage) in 38 primary influent sewage samples collected from two wastewater treatment facilities [34]. The boundary of each box closest to zero indicates the 75^th^ percentile. Whiskers above and below each box indicate the 10^th^ and 90^th^ percentiles. A black circle denotes outlier measurements.

### 3.5 Comparison of culture-based GB-124 with ΦB124-14 and crAssphage bacteriophage-like qPCR genetic markers

Previously reported data and sample material from 38 primary influent sewage samples [34] was used to further evaluate trends between novel ΦB124-14 and crAssphage bacteriophage-like target sequence occurrence and assess potential correlations with culture-based GB-124 paired measurements. Due to the strong correlation between CPQ_056 and CPQ_064 crAssphage-like qPCR data from geographically dispersed primary influent sewage samples (r = 0.988; p < 0.001), CPQ_056 was selected to estimate crAssphage bacteriophage-like target sequence concentrations. The larger test quantities (2 mL sewage per reaction) from 10 L primary influent samples yielded measurements in the range of quantification for all qPCR assays and sewage samples tested (**Figure 2**). Pearson product moment correlation analyses between paired measurements indicates a strong agreement between PS28 and PS30 (r = 0.938; p < 0.001) and a moderate agreement between ΦB124-14 and crAssphage bacteriophage-like target sequences (PS28 versus CPQ_056: r =0.593, p < 0.001; PS30 versus CPQ_056: r = 0.646, p < 0.001). However, ΦB124-14 and crAssphage bacteriophage-like genetic measurements showed no significant correlations with culture-based GB-124 paired measurements (r ≤ 0.221, p ≥ 0.183).

### 3.6 Experimental controls

A series of controls were used to identify high-quality qPCR data. Calibration model performance parameters are shown in **Table 2**. A total of 43 outliers (6.7%) were observed (645 total measurements) with 97.7% corresponding to the lowest standard concentration used as template. Extraneous DNA control reactions indicated no evidence of contamination (*n* = 375 reactions). All DNA preparations exhibited no amplification inhibition in duplex HF183/BacR287 experiments (*n* = 184 samples). IAC acceptance thresholds ranged from 32.9 C_q_ to 33.6 C_q_ and competition thresholds ranged from 26.0 C_q_ to 26.2 C_q_ for HF183/BacR287. Instrument run-specific HF183/BacR287 IAC proficiency testing yielded a 100% pass rate with NTC VIC C_q_ standard deviations ranging from 0.10 to 0.27 (acceptance criteria ≤ 1.16) [40].

## 4. DISCUSSION

### 4.1 Identification of human-associated ΦB124-14 bacteriophage-like genetic regions

The complete genome of a human gut-associated *Bacteroides* bacteriophage, designated as ΦB124-14 was recently reported [20]. ΦB124-14 was originally isolated from a wastewater sample collected from a treatment facility in the United Kingdom and is propagated on *Bacteroides* sp. GB-124 [41]. Ecogenomic sequence analyses suggest that portions of the ΦB124-14 genome are routinely found in sewage metagenomic DNA libraries [21]. To investigate the potential use of select ΦB124-14 genetic regions for sewage-associated qPCR method development, we employed a bioinformatic-based “biased genome shotgun strategy” [9] combined with high-throughput laboratory testing to identify putative sewage-associated genetic regions. Fifty-three primer sets targeting 29 ORFs covering 6,231 base pairs were subject to three rounds of performance testing with each round introducing more stringent criteria. Approximately 72% of candidate primer sets generated expected end-point PCR products in primary influent sewage samples, supporting bioinformatic predictions that ΦB124-14 is commonly found in wastewaters. The remaining 28.3% of candidate primer sets were not detected in sewage samples. No PCR amplification from these primer sets may have been due to sequence variation at primer hybridization sites, lack of assay optimization, or low genetic conservation between individuals resulting in low template availability.

Almost 80% of candidate primer sets in round one testing did not yield amplification products of the expected size in non-human sources used as template DNA, supporting the bioinformatic prediction that ΦB124-14 is closely associated with human sewage [21]. While limited, the occurrence of false positives with some primer sets suggests that portions of the ΦB124-14 genome share homology with microorganisms associated with the gut of other animals tested in this study. Previous research suggests that ΦB124-14 predicted ORF encoding regions should be treated in an independent fashion when investigating host association due to the mosaic architecture and plasticity of bacteriophage genomes [21, 42, 43]. Findings reported here support this assertion where 33.3% (10 of 30) of ORF genetic regions targeted by candidate primer sets elicited a false positive detection in round one screening.

After three rounds of screening, PS28 and PS30 primer sets were selected for qPCR method development. These genetic regions represent the most abundant and sewage-associated candidate DNA targets based on end-point PCR amplification conditions and reference fecal and sewage collections used in this study. Both primer sets target the forward strand of the ΦB124-14 genome identified as ORF 46 and are predicted to encode for a hypothetical protein [20]. Candidate end-point PCR primer sets PS29 and PS31 also targeted the ORF 46 genetic region and did not yield an amplification product of the expected size when non-human sources were used as DNA template in round one of screening but failed due to the absence of a PCR amplification product of the expected size when primary influent sewage was used as DNA template (PS29) or yielded a spurious amplification product (PS31). Because this study did not employ optimization of end-point PCR amplification conditions (i.e., annealing temperature), it is possible that PS29 and PS31 primer sets could pass screening criteria with additional optimization.

DNA sequence verification of amplicons from primary influent sewage samples confirmed amplification of expected ΦB124-14 bacteriophage-like sequences for PS28 and PS30. However, sequencing results demonstrate that different regions of the same ORF can exhibit variable degrees of sequence homology. For example, the PS30 sequence library yielded a single ASV consisting of 99.9% of total processed sequence reads, inferring a potentially high level of genetic stability for this marker. In contrast, PS28 yielded 16 ASV types where the most dominant ASV comprised 81.4% of total processed sequence reads (PS28_ASV_1, **Figure 1, Panel B**). In this study, only ORF encoding regions were interrogated to minimize genetic regions with high recombination or mutagenic potential. While DNA sequencing suggests some level of sequence stability, novel ΦB124-14 bacteriophage-like qPCR assays should be monitored in the future to confirm amplification of intended targets, especially for the PS30 qPCR assay.

### 4.2 Performance of ΦB124-14 bacteriophage-like qPCR assays

The performance of novel PS28 and PS30 ΦB124-14 bacteriophage-like qPCR assays was evaluated through a series of paired experiments with established DNA-based bacterial (HF183/BacR287 and HumM2) and viral (CPQ_056 and CPQ_064) qPCR assays. Standard calibration models indicated that all qPCR assays are highly optimized exhibiting amplification efficiencies ranging from 0.90 to 1.03 and R^2^ ≥ 0.997. All qPCR assays exhibited specificities ≥ 92% and sensitivities ≥ 86.1%, exceeding the expert consensus 80% benchmark metric for water quality applications [27]. Both ΦB124-14 bacteriophage-like qPCR assays exhibited 100% specificity indicating that the addition of a hydrolysis probe to the PS28 end-point PCR primer set eliminated low level cross-reactivity with horse, chicken, and cat sources observed in round three screening. In addition to superior specificity, ΦB124-14 bacteriophage-like qPCR assays exhibited 86.1% sensitivity to 36 geographically dispersed primary influent sewage samples collected across the continental United States when 1 ng of total DNA was used as template. However, other bacterial and viral human-associated assays exhibited 100% sensitivity. A reduced detection level with ΦB124-14 bacteriophage-like qPCR assays could suggest that these genetic markers may not be shed at high concentrations in United States populations. This assertion aligns with previous studies investigating the geographic occurrence of cultured GB-124 where reported concentrations ranged from 2 to 4.7 log_10_ plaque forming units (PFU) per 100 mL in final influent from the United Kingdom [18] but were only 0.16 to 2.2 log_10_ PFU per 100 mL in primary sewage effluent from the United States [14]. In addition, ΦB124-14 sequences were reported to share homology with 83.8% of individual human gut microbiomes from the United Kingdom and none from the Americas [20]. Additional research on the occurrence of ΦB124-14 bacteriophage-like qPCR assay genetic markers in sewage from different geographic locales is needed to better ascertain the sensitivity of these novel viral assays.

The HF183/BacR287 qPCR assay was the top performing bacterial method using sewage and fecal reference samples from this study, yielding a specificity of 98% and 100% sensitivity, providing further evidence that this is an effective human-associated fecal source identification tool [27, 44–46]. CrAssphage-like and HF183/BacR287 genetic markers occurred at concentrations up to 724 times higher compared to PS28 and PS30 levels in primary influent sewage samples, on average. CPQ_056 and CPQ_064 were the only qPCR assays to elicit false positives within the range of quantification when non-human DNA was used as template with all instances originating from domestic cat fecal samples. Cross-reactivity of these crAssphage-like qPCR assays has also been reported in cat fecal samples from Australia [47] and could confound future fecal source identification studies where feline waste is a plausible source of fecal pollution.

### 4.3 ΦB124-14 and crAssphage bacteriophage-like qPCR assay genetic marker trends with cultivated GB-124 in sewage

A significant limitation for traditional PCR-based detection of viral targets in wastewater and ambient water sample types is the inability to determine infectivity status. As a result, there is a need for viral indicators where protocols are available to detect by both cultivation and genetic methodologies. To evaluate potential trends of ΦB124-14 and crAssphage bacteriophage-like genetic marker occurrence compared to GB-124, paired measurements were investigated from 38 primary influent sewage samples using archived nucleic acid material and corresponding GB-124 measurements from a previously reported study [34]. Findings demonstrate that ΦB124-14 bacteriophage-like genetic markers can be consistently measured within the range of quantification when larger volumes of sewage are utilized resulting in a higher template quantity (**Figure 2**) compared to the standardized 1 ng of total DNA quantity used for performance assessment. ΦB124-14 bacteriophage-like qPCR assay paired measurements showed strong agreement (r = 0.938) which is expected because both assays target different parts of the same genetic region (ORF 46). CrAssphage bacteriophage-like genetic marker levels were also significantly correlated with paired ΦB124-14 bacteriophage-like measurements albeit to a lower degree (r ≥ 0.593, p < 0.001), suggesting that these different types of bacteriophages likely exhibit minor variations in human waste occurrence levels. However, both ΦB124-14 and crAssphage bacteriophage-like measurements showed no significant correlation with culture-based GB-124 (r ≤ 0.221, p ≥ 0.183). The lack of agreement between ΦB124-14 bacteriophage-like genetic markers and GB-124 paired measurements in untreated sewage samples could arise for several reasons. For example, ΦB124-14 is reported to exhibit a narrow host range of closely related *Bacteroides* spp. including the *B. fragilis* GB-124 host strain [20], potentially due to extreme niche specialization [48] leading to a highly specific bacterial host interaction. It is possible that GB-124 may be infected by other bacteriophage strains closely related to ΦB124-14, confounding a potential agreement between measurements. In addition, it is important to note that GB-124 was isolated from a wastewater sample in 2007. Over 15 years later, the same isolate has subsequently been used across all peer-reviewed published experiments to date. Repeated propagation from the original stock preparation could lead to the accumulation of mutations that could potentially influence the interaction between ΦB124-14 and the current GB-124 host preparation [49]. Additional research is required to elucidate the mechanisms accounting for the lack of agreement between ΦB124-14 bacteriophage-like qPCR assay and GB-124 paired measurements.

### 4.4 Potential applications for ΦB124-14-like Bacteroides bacteriophage qPCR assays

Even though the novel ΦB124-14 bacteriophage-like genetic markers measured by PS28 and PS30 qPCR assays exhibited poor agreement with GB-124 measurements in primary influent sewage, these assays exhibited a strong association with sewage (100% specificity) and were detected in most primary influent sewage samples collected across the United States (> 86% of samples). However, PS28 and PS30 genetic marker concentrations were substantially lower compared to top performing DNA-based bacterial (HF183/BacR287 and HumM2) and viral (CPQ_056 and CPQ_064) human-associated qPCR assays. As a result, ΦB124-14 bacteriophage-like qPCR assays may have limited utility in surface water fecal source identification applications in the United States, especially when only trace levels of sewage are present. However, when sewage was tested at higher concentrations, ΦB124-14 bacteriophage-like sequences were routinely detected within the range of quantification suggesting that these genetic markers may prove valuable for wastewater treatment efficacy applications. Because both ΦB124-14 and GB-124 occurrence can vary by geographic location and multiple studies report that GB-124 levels are higher in European compared to other populations [8, 14, 18, 20], additional research is needed to investigate the performance of ΦB124-14 bacteriophage-like qPCR assays in other locales, especially in Europe.

## 5. CONCLUSIONS

A bioinformatic “biased genome shotgun approach” was combined with intensive laboratory testing to interrogate the ΦB124-14 genome to develop two novel bacteriophage-like qPCR methods for the characterization of human sewage in wastewater and surface waters. Key findings included:

- ΦB124-14 bacteriophage-like qPCR assays exhibited superior specificity compared to top performing DNA-based bacterial (HF183/BacR287 and HumM2) and viral (CPQ_056 and CPQ_064) human-associated fecal source identification methods.
- ΦB124-14 bacteriophage-like sensitivity exceeded the expert consensus benchmark of 80% [27] but occurred at substantially lower concentrations compared to other commonly used human-associated fecal source identification methods in continental United States sewage samples.
- Performance assessment of ΦB124-14 bacteriophage-like qPCR assays with sewage samples collected from other geographic locales outside of the United States may yield more favorable results, especially from European populations.
- Future studies may be needed to verify the temporal genetic stability of ΦB124-14 bacteriophage-like qPCR assays even though results in this study suggest some level of conservation.
- Paired measurements of ΦB124-14 bacteriophage-like genetic markers and culture-based GB-124 in sewage samples from the United States demonstrated poor agreement. The exact mechanisms responsible remains unknown.
- Novel ΦB124-14 bacteriophage-like qPCR assays represent a potentially valuable addition to the sewage identification toolbox, providing the means to quantify sewage pollution levels and compare occurrence with a culture-based virus method.

## Data Availability

All data produced in the present study are available upon reasonable request to the authors

## Acknowledgements

Information has been subjected to U.S. EPA peer and administrative review and has been approved for external publication. Any opinions expressed in this paper are those of the authors and do not necessarily reflect the official positions and policies of the U.S. EPA. Any mention of trade names or commercial products does not constitute endorsement or recommendation for use. Amplicon sequence data were processed using resources from the Minnesota Supercomputing Institute.

