## Supplemental Information for "ΦB124-14 *Bacteroides* Bacteriophage-Like Quantitative Real-Time PCR Assays for Human Sewage Pollution Measurement"

**List of Tables:**

**Table S1:** Summary of reference fecal sample collection.
**Table S2:** Candidate primer set sequence information and results of end-point PCR primer set testing for rounds I, II, and III.

**Table S1:** Summary of reference fecal sample collection.

| Host Information | | Location | Count |
|---|---|---|---|
| *Common Name* | *Scientific Name* | | |
| Cat | *Felis catus* | Cincinnati, OH | 10 |
| Calf | *Bos taurus* | Tillamook, OR | 10 |
| Cow | | | 10 |
| Dog | *Canis lupus familiaris* | Cincinnati, OH | 10 |
| Chicken | *Gallus gallus domesticus* | Cincinnati, OH | 10 |
| Goose | *Anser anser domesticus* | Cincinnati, OH | 10 |
| Horse | *Equus caballus* | | 10 |
| Raccoon | *Procyon lotor* | Boulder, CO | 10 |
| Pig | *Sus scrofa* | Middletown, OH | 10 |
| Seal | *Halichoerus grypus* | Cape Cod, MA | 10 |

**Table S2:** Candidate primer set sequence information and results of end-point PCR primer set testing for rounds I, II, and III.

| Primer Set | Primer Sequence 5' → 3' | ORF | Round I | Round II | Round III | Reason for Failure |
|---|---|---|---|---|---|---|
| 1 | F: AGAAAACCCACCGATTGAGGA<br>R: TCCTTTGCCTGTTCGTTAGTTT | 1 | Fail | NT | NT | Spurious bands |
| 2 | F: AGGGATGTAGGGTAAAGAAAGAACC<br>R: AGCCTCTTTTATATACCCACGCTT | 2 | Fail | NT | NT | Spurious bands |
| 3 | F: GGAGGCAGCAAGGAAGAGAA<br>R: GGGATCGTAGGAGACATGCAC | 17 | Fail | NT | NT | Spurious bands |
| 4 | F: TAATACGCCAACACCGGAAGT<br>R: CGGATGTTTTAGAGCCGCCA | 17 | Fail | NT | NT | Spurious bands |
| 5 | F: GCCGTTCTATCGGCAAAGAC<br>R: GCGTTCTCTCCCATCAACCT | 18 | Pass | Fail | NT | Spurious bands in cow and pig samples |
| 6 | F: ACCGCCCTTGTGTTCATGTT<br>R: TGAACCTACGCACACCATTCA | 18 | Pass | Fail | NT | False positive to dog sample |
| 7 | F: TCGACAGAGCCGGACAAGTA<br>R: CTACCGGAAGTTCCAGGCATT | 19 | Fail | NT | NT | Spurious bands |
| 8 | F: GCGCACATAAATTCGGCAGG<br>R: CTTGCTGTAAGCTCGCCTGT | 20 | Fail | NT | NT | Spurious bands |
| 9 | F: TCTTGGTTGTCGGTGGGTAA<br>R: CGTCCTCTGTGAGATACGCT | 20 | Fail | NT | NT | Spurious bands |
| 10 | F: GGGTCTCAAAGTACAACGCC<br>R: CTCCAATTGTTGTCGGGGGT | 20 | Fail | NT | NT | Spurious bands |
| 11 | F: ACGTACACTAAATTGGATGGCA<br>R: TTTCCAAGTGAAGGAAACCGA | 21 | Fail | NT | NT | Spurious bands |
| 12 | F: TAAGGGTATCGGTGTTGGTGC<br>R: TTGCCTCCTTTTTGGCGGTA | 22 | Fail | NT | NT | Spurious bands |
| 13 | F: AGATTGGTGTGCTGACTGTGG<br>R: TATGCGCAATTAACGAACGCC | 27 | Fail | NT | NT | Spurious bands |
| 14 | F: CCGATCACAGCAAACGAAGT<br>R: GCACAGATATGCTTTACACATGCT | 28 | Fail | NT | NT | Spurious bands |
| 15 | F: ACTAAAGGCGAGAGAGGGGATA<br>R: GAACCAACACGAAGTGCAACA | 29 | Fail | NT | NT | Spurious bands |
| 16 | F: AATGCACCCTTGGACGTATG<br>R: AGGATGAACTTTCCCGGATCTC | 29 | Fail | NT | NT | Spurious bands |
| 17 | F: TGACCTAAAGGTAGGCGAGGA<br>R: ACCCATTTCCCCAAAGAAGCA | 30 | Fail | NT | NT | Spurious bands |
| 18 | F: ACCCCCAAAGGTGATAAGGTT<br>R: TCTTTTGTTTCACGCTCCGGT | 30 | Fail | NT | NT | Spurious bands |
| 19 | F: ATGCAATTTACCGGAGCGTG<br>R: TCTCGTTACTACGCATTCTCTCC | 30 | Fail | NT | NT | Spurious bands |
| 20 | F: TTCTTTGCGGAGGTACCGAA<br>R: TTCTCGCATGCGTCTCCATT | 31 | Fail | NT | NT | Spurious bands |
| 21 | F: GAGACGCATGCGAGAAATGT<br>R: ATTTCCCCATCTCTCACCCTC | 31 | Fail | NT | NT | Spurious bands |
| 22 | F: CGGTAGCGGAAACTATACCGT<br>R: ACCGCAATTACGTTCTACGC | 32 | Fail | NT | NT | Spurious bands |
| 23 | F: CCCGGCTGTTATTGTTATACTGAT<br>R: GAAAACGGCAACGAAAGAGCC | 33 | Fail | NT | NT | Spurious bands |
| 24 | F: ACGGAAAAGAGATAAGCGAACA<br>R: TCGCCCAAAGAGAAAGATCG | 34 | Fail | NT | NT | Spurious bands |
| 25 | F: GTGCATGGTTAGGAGATGGTGA<br>R: AACGCCTCGTTGTACTTCCTT | 43 | Fail | NT | NT | Spurious bands |
| 26 | F: GGCAGATTCGGACAACCTGT<br>R: AGCCTTTCTAATGGTCTACCGTG | 43 | Fail | NT | NT | Spurious bands |
| 27 | F: AGGGGAGAGAAATAAGAGAGGCT<br>R: ACCATCTCCTAACCATGCACC | 43 | Fail | NT | NT | Spurious bands |
| 28 | F: AGCGAATGGGCTACTCTGAAC<br>R: TGCCTCAATCGCTTTACCCAA | 46 | Pass | Pass | Pass | --- |

| | | | | | | |
|---|---|---|---|---|---|---|
| 29 | F: GGTGTATGGACGCAGTAGGA<br>R: TCTGTTCAGAGTAGCCCATTCG | 46 | Fail | NT | NT | Spurious bands |
| 30 | F: GCGGAGAGAAAAGGACGTTA<br>R: TCCTACTGCGTCCATACACC | 46 | Pass | Pass | Pass | --- |
| 31 | F: GTGCAGGGCAACATGAAAGAAT<br>R: CCTCCATTGCCTCGCACATC | 46 | Fail | NT | NT | Spurious bands |
| 32 | F: ATTGAGAAGCGCACAGAAGC<br>R: TCATCTACTTTCTTTTCCGCCT | 58 | Fail | NT | NT | Spurious bands |
| 33 | F: GTTCCGGTAGCCCTGCAATA<br>R: CGAGCGCACAACATACAAGG | 59 | Fail | NT | NT | Spurious bands |
| 34 | F: CCTTTGTTCCTATGTGCGCC<br>R: CGTTTGCAAGTGGGAATTGCT | 59 | Fail | NT | NT | Spurious bands |
| 35 | F: CCGGAGGCGGGAAAGAATTTA<br>R: CCACTTTGCCACGAAATAGGC | 59 | Fail | NT | NT | Spurious bands |
| 36 | F: GCGATGAAGCAATTCCCACTT<br>R: CGTGCTGTTTTCGTTCTGCAT | 59 | Pass | Fail | NT | Spurious bands in dog sample |
| 37 | F: GGCTTACAAAGCCTTCCGATA<br>R: CAGAGTATAACATCTCTTTCCCTCC | 60 | Pass | Fail | NT | Spurious bands in cow and dog samples |
| 38 | F: TCCGGGACGCAGGAAGTAT<br>R: TTCCCCGACAGCAAGTAACC | 61 | Fail | NT | NT | Spurious bands |
| 39 | F: TGCGTACATGGAACGATTGG<br>R: AACTGCTTCGTGGTCTATCCC | 61 | Fail | NT | NT | Spurious bands |
| 40 | F: AAGTGGCAACTTCGGACAGC<br>R: CGTCTGTACATTCGGACACC | 61 | Fail | NT | NT | Spurious bands |
| 41 | F: AGCAAACCTAAACGAGATTGCG<br>R: CCGTCTTTTCCCGTAGCCAT | 62 | Fail | NT | NT | Spurious bands |
| 42 | F: GGCTATTCTGAGCCGATTTTGC<br>R: CTTCGCGCGTGGCTATACTA | 62 | Fail | NT | NT | Spurious bands |
| 43 | F: ATAGTATAGCCACGCGCGAA<br>R: GACTCGAACGGGAATGTTAGC | 62 | Pass | Fail | NT | Spurious bands in dog, cow, and pig samples |
| 44 | F: AGCGCATACGAGAAGCTAATCA<br>R: CCTTTCGGGGTAACAACCGT | 63 | Fail | NT | NT | Spurious bands |
| 45 | F: GATGCAGCAATCGCTTCCAA<br>R: CTACTCGCAGAGAGCTACCC | 64 | Fail | NT | NT | Spurious bands |
| 46 | F: GTGAACTTTCACGTCTTGGGC<br>R: CCACACCTTGCATCCCTTTG | 65 | Fail | NT | NT | Spurious bands |
| 47 | F: CAAAGGGATGCAAGGTGTGG<br>R: TCAGCTGACAATCCGAAAGCA | 65 | Fail | NT | NT | Spurious bands |
| 48 | F: ACACAAAACGACAGCTTGGA<br>R: AGAGATTACGCCCAAGACGTG | 65 | Fail | NT | NT | Spurious bands |
| 49 | F: TGAACGCCCTACGAGGAACT<br>R: TCCCGCTATACGACAGGTTC | 66 | Fail | NT | NT | Spurious bands |
| 50 | F: TGGGCGTTTATCATTCAGCCT<br>R: CCACCAGCATGTTTACGCTTT | 66 | Fail | NT | NT | Spurious bands |
| 51 | F: TTACCGGAGAAAAGGCGCTG<br>R: CTGTTCGGCAGCAAGCAATAA | 67 | Fail | NT | NT | Spurious bands |
| 52 | F: GCCGAAAAAATCGGCTGTACTA<br>R: TTCTCCGGTAAAGCCCATCG | 67 | Fail | NT | NT | Spurious bands |
| 53 | F: ACAGTACCTTGGATATGGAGGC<br>R: TCCGTTCTTTGTACTCCGCA | 68 | Fail | NT | NT | Spurious bands |

RF denotes the open reading frame.
NT indicates the primer set was not tested.
